# Methodological quality of recommendations on vitamin D and calcium – a systematic review of bone health guidelines

**DOI:** 10.1101/2021.05.01.21256288

**Authors:** Zhaoli Dai, Joanne E McKenzie, Sally McDonald, Liora Baram, Matthew J Page, Margaret Allman-Farinelli, David Raubenheimer, Lisa A. Bero

## Abstract

There are numerous guidelines developed for bone health. Yet it is unclear the differences in guideline development methods explain the variability in recommendations for vitamin D and calcium intakes. The objective of this systematic review was to collate and compare recommendations for vitamin D and calcium across bone health guidelines, assess methods used to form the recommendations, and explore which methodological factors were associated with these guideline recommendations. We searched MEDLINE, EMBASE, CINAHL and other databases indexing guidelines to identify records in English between 2009 and 2019. Guidelines or policy statements on bone health or osteoporosis prevention for generally healthy adults aged ≥40 years were eligible for inclusion. Two reviewers independently extracted recommendations on daily vitamin D and calcium intake, supplement use, serum 25 hydroxy vitamin D [25(OH)D] level, and sunlight exposure; assessed guideline development methods against 25 recommended criteria in the World Health Organization (WHO) Handbook for Guideline Development; and, identified types of evidence underpinning the recommendations. We included 47 eligible guidelines from 733 records: 74% of the guidelines provided vitamin D (200∼600-4000 IU/day) and 70% provided calcium (600-1200 mg/day) recommendations; 96% and 88% recommended vitamin D and calcium supplements, respectively; and 70% recommended a specific 25(OH)D concentration. The mean of meeting 25 WHO methodological criteria per guideline was 10 (95% CI: 9-12; interquartile range: 6-15). There was uncertainty in the associations between the methodological criteria and the proportion of guidelines that provided recommendations on daily vitamin D or calcium. Various types of evidence, ranging from previous bone guidelines, nutrient reference reports, systematic reviews, observational studies, to perspectives/editorials were used to underpin the recommendations. In conclusion, there is considerable variability in vitamin D and calcium recommendation and in guideline development methods in bone health guidelines. Effort is required to strengthen methodological rigor of guideline development and utilize the best available evidence to underpin public health nutrition.

**Highlights:**

- This systematic review provides evidence on the variabilities in vitamin D and calcium recommendations as well as guideline development methods in 47 bone health guidelines globally.
- Our findings point to continued effort to utilize the best available evidence to underpin nutrition recommendations and strengthen methodological rigor of guideline development in bone health guidelines.

**Registration of protocol:** PROSPERO: CRD42019126452

## Introduction

Due to global aging, the prevalence and incidence of osteoporosis and fractures continue to rise in both developed and developing countries (1). The social and economic burdens associated with osteoporosis, particularly fractures at the hip, are substantial, including disability, fracture recurrence, and premature mortality (2-4). However, an effective and feasible non-pharmacological prevention strategy is yet to be widely endorsed.

Vitamin D and calcium are two essential nutrients for normal bone growth and bone maintenance. Calcium plays a crucial role in skeletal mineralization to support bone strength and muscle contraction. Vitamin D facilitates calcium absorption via its active hormonal form, 1,25-dihydroxycholecalciferol [1,25(OH)2D3], working together with parathyroid hormone to maintain calcium homeostasis (1, 5). There is no doubt that maintaining sufficient vitamin D and calcium is vital at every age. However, factors such as sunlight exposure and dietary habits, genetic and cultural backgrounds, and the aging process can contribute to different physiological needs for vitamin D and calcium (6-9). To date, what is considered the appropriate level of vitamin D and calcium supplementation as well as serum 25-hydroxycholecalciferol [25(OH)D] concentration remains controversial for bone health (10).

In public health and clinical guidelines on bone health, vitamin D and calcium recommendations constitute an important non-pharmacological strategy for bone maintenance and the prevention of osteoporosis and fractures. However, specific recommendations vary in guidelines from different countries and even differ among organizations within a country. This is partially due to the conflicting evidence seen in the effectiveness of vitamin D and calcium supplements on bone mineral density (11, 12) and fractures (13-16). Furthermore, using these supplements at high doses may pose adverse events, including falls (17, 18), cardiovascular diseases, and kidney stones (16, 19).

Development of guideline recommendations should be informed not only by the synthesis of the available evidence but also by stakeholders’ representation and perspectives on intervention strategies, as well as considerations of health equity, acceptability, and feasibility. Furthermore, commercial influence should be minimized when formulating the recommendations (20). It is currently unclear to what extent the variability in vitamin D and calcium recommendations is related to the methods used to develop the bone health guidelines.

The objective of this study was to collate and compare recommendations for vitamin D and calcium across bone health guidelines, appraise the methodological quality of the guideline recommendations, and identify methodological factors that might affect the recommendations of vitamin D and calcium on daily intake, dietary and supplemental intake, serum level of 25(OH)D, and sunlight exposure.

## Methods

We registered this systematic review in PROSPERO (registration number: CRD42019126452) in March 2019 and published a peer-reviewed protocol (21).

### Data sources and searches

Working with an experienced academic librarian, we searched for bone health guidelines or policy statements in the following electronic databases: MEDLINE (via OVID), EMBASE (via OVID), CINAHL (via EBSCO), Practice-Based Evidence in Nutrition, National Guideline Clearinghouse (by Agency for Healthcare Research and Quality, AHRQ), National Institute for Health and Care Excellence (NICE), and Guidelines International Network (GIN) in March 2019. The search period was restricted to 1 January 2009 until 28 February 2019. For other databases, the searches were based on a single keyword or a combination of vitamin D (or calcium), bone, osteoporosis, guideline/policy, and recommendation as search terms. The search strategy used to retrieve the guidelines in MEDLINE, EMBASE, and CINAHL as well as the other databases indexing guidelines mentioned above are described in the **Supplemental Materials**. Additionally, we searched the website of the International Osteoporosis Foundation to capture missing guidelines or policy statements.

### Study (guideline/policy statement) selection

Details of the inclusion and exclusion criteria were published in our protocol (21). Briefly, we only included the most up-to-date bone health guidelines developed by nationally or internationally recognized government authority, or a medical/academic society or organization within a country or a special region (such as Hong Kong or Taiwan). Our target populations were generally healthy adults aged 40 years and older who were at risk of developing osteoporosis. The reason for selecting this age group was that some women might experience early menopause as young as 40 years (22, 23). Due to the available resources, we only included guidelines written in English. We excluded bone health guidelines related to the management of osteoporosis (such as postmenopausal women under a physician’s care), secondary osteoporosis (e.g., osteoporosis due to rheumatoid arthritis) or for a particular group of population or those with health condition (21). Government reports on nutrient reference values (nutrient requirements) were not included in this review, as they use rigorous methodology, including systemic reviews, dose-dependent, factorial approach, or balance study, and consideration of characteristics of dietary intake and other considerations for the target population to determine dietary reference intakes (24, 25). As the guiding principles of nutrient intakes for public health policy, these reports are out of scope when we are interested in the methodological rigor of public health guideline development process.

One reviewer (ZD) screened titles and abstracts of all the retrieved records. Full text of potentially eligible guidelines or policy statements was further assessed and finalized among three reviewers (ZD, SM and LB) based on reading the full text thoroughly and the eligibility criteria noted above.

### Data extraction and quality assessment

Data on guideline characteristics, vitamin D and calcium recommendations, and evidence cited to support the recommendations were extracted (**Table 1**). For both nutrients, the extraction of daily vitamin D [(international unit (IU)/day or µg/day] and calcium (mg/day) recommendations were extracted as they were reported in the guidelines.

**Table 1.** Data items extracted from guidelines

| **Characteristics of guidelines** | **Recommendations on vitamin D and calcium** | **Evidence cited to support recommendations** |
| --- | --- | --- |
| Title; | Daily intake of vitamin D (IU/day); | Verbatim text that referenced the supporting studies / guidelines; |
| Guideline developing authority or organization; | Dietary intake of vitamin D rich foods; | Full text articles of supporting studies grouped to the following types [Previous guideline from other countries; international guidelines (e.g., guidelines published from the World Health Organization; Systematic review of RCTs; Systematic review of non-randomized studies; Clinical trial; Cohort study; Cross-sectional study; Case-control study; Other (such as narrative review, editorial, and commentary)] |
| Publication year; | Dosage of vitamin D supplements (IU/day); |  |
| Age group; | Daily intake of calcium (mg/day); |  |
| Sex of target population; | Dietary intake of calcium rich foods; |  |
| Funding source | Dosage of calcium supplements (mg/day); |  |
|  | Sunlight exposure; |  |
|  | Recommended level of serum 25(OH)D (nmol/L) |  |

To ensure consistency of extractions of the recommendations – which comprised quantitative (i.e. with numerical values) and qualitative (i.e. descriptive text) information – we adopted the criteria proposed by Woolf and colleagues (26). To evidence underpinning the recommendations, details for the cited evidence was extracted. This was followed by retrieval of the full text, which was then categorized by evidence type. Additionally, more than one evidence type could be selected per recommendation. Two reviewers (ZD and SM or LB) independently extracted the data and any discrepancies in the data extraction or categorizations were resolved via discussion or through consultation with the senior author (LAB).

### Assessment of guideline development methods

Using a content analysis approach, two reviewers (ZD and SM or LB) independently appraised each guideline according to a set of 25 criteria adopted from the 2014 WHO Handbook for Guideline Development (27). The WHO methodological criteria include those in the Appraisal of Guidelines for Research and Evaluation (AGREE) II (28) that are commonly used for assessing guideline quality, and also include additional considerations such as using systematic review methods to search, retrieve, and synthesize evidence, transparency of types of evidence used, rating of the importance of the health outcome, and consideration of health equity, acceptability, and feasibility in the process of formulating recommendations (27). Details of the criteria and our rationale for using the WHO guideline methods were described in our protocol (21).

For each guideline, we rated whether each of the recommended WHO methodological criteria was applied (**Supplemental Table 1**), using the response options Yes, No, or Unclear. If a method was used (response option “Yes”), verbatim text from the guideline (or supplementary materials) was extracted. “No” was selected when a guideline explicitly stated that it did not adopt the specific process (e.g., “there was no stakeholder involvement in this guideline.”). If the process was not explicitly stated as not being used or was not described, we used the response option “Unclear”. Any discrepancies for appraisal of the guideline development processes were resolved by discussion among the reviewers or by consultation with the senior author (LAB).

All data extraction and methods assessment of guideline development processes were captured and stored using the Research Electronic Data Capture, an electronic data capture tool hosted at the University of Sydney (29). The data were then exported to Excel for data cleaning and analysis in other statistical programs.

### Data synthesis and analysis

Descriptive summary statistics using frequencies and percentages were used to summarize guideline characteristics and the recommendations made for daily recommended intake of vitamin D (IU/day; 1μg = 40 IU), calcium (mg/day), and recommended level of serum 25(OH)D (1 ng/ml = 2.5 nmol/L). We also recoded some of the recommendations into dietary recommendation made for either vitamin D/calcium-rich food (Yes/No), supplement recommendation made for either vitamin D/calcium supplements (Yes/No), and sunlight exposure recommendation (Yes/No). For the types of evidence supporting the recommendations, we combined three groups of guidelines [Source country of guideline, Previous guideline from other countries, and International guidelines (e.g., guidelines published from the World Health Organization)] into one category.

For assessing guideline development methods, we calculated the mean (95% confidence interval, CI) of WHO methodological criteria met for each guideline and the proportion (95% CI) of guidelines that fulfilled each of the criteria. We examined the association between each of the WHO methodological criteria and forming recommendations of daily intake for vitamin D and calcium to investigate whether variability in the quality of the guideline methods could potentially explain variations in the recommendations. Because of the small sample of guidelines, we combined different types of recommendations into a binary variable, i.e., vitamin D or calcium recommendation made (or not made). We calculated risk difference (RD) between a guideline process had met or not met a WHO guideline development criterion and the recommendations made on vitamin D / calcium (e.g., compared guidelines that met “discipline representation” with those that did not, what was the difference for guidelines where a recommendation was made for daily intake of vitamin D/calcium). We used a confidence level of 99% in the calculation of the confidence intervals for these differences, rather than 95%, to facilitate greater caution in our interpretation of the results due to the number of associations examined. We calculated Fisher’s exact P-value because there were a small number of guidelines.

All analyses were performed using SAS (version 9.4) or Stata version 16 (30). The forest plot was produced using the metaprop package (31). A two-sided p-value (<0.05) is considered statistically significant.

## Results

### Results of search

After removal of 160 duplicates from 733 records identified from different databases, 573 records remained. A further 456 records were removed after screening title or abstract, and 117 records underwent full-text screening, from which, 47 eligible guidelines were included (**Figure 1**). **Supplemental Table 2** lists the guidelines included in this review.

**Figure 1.**
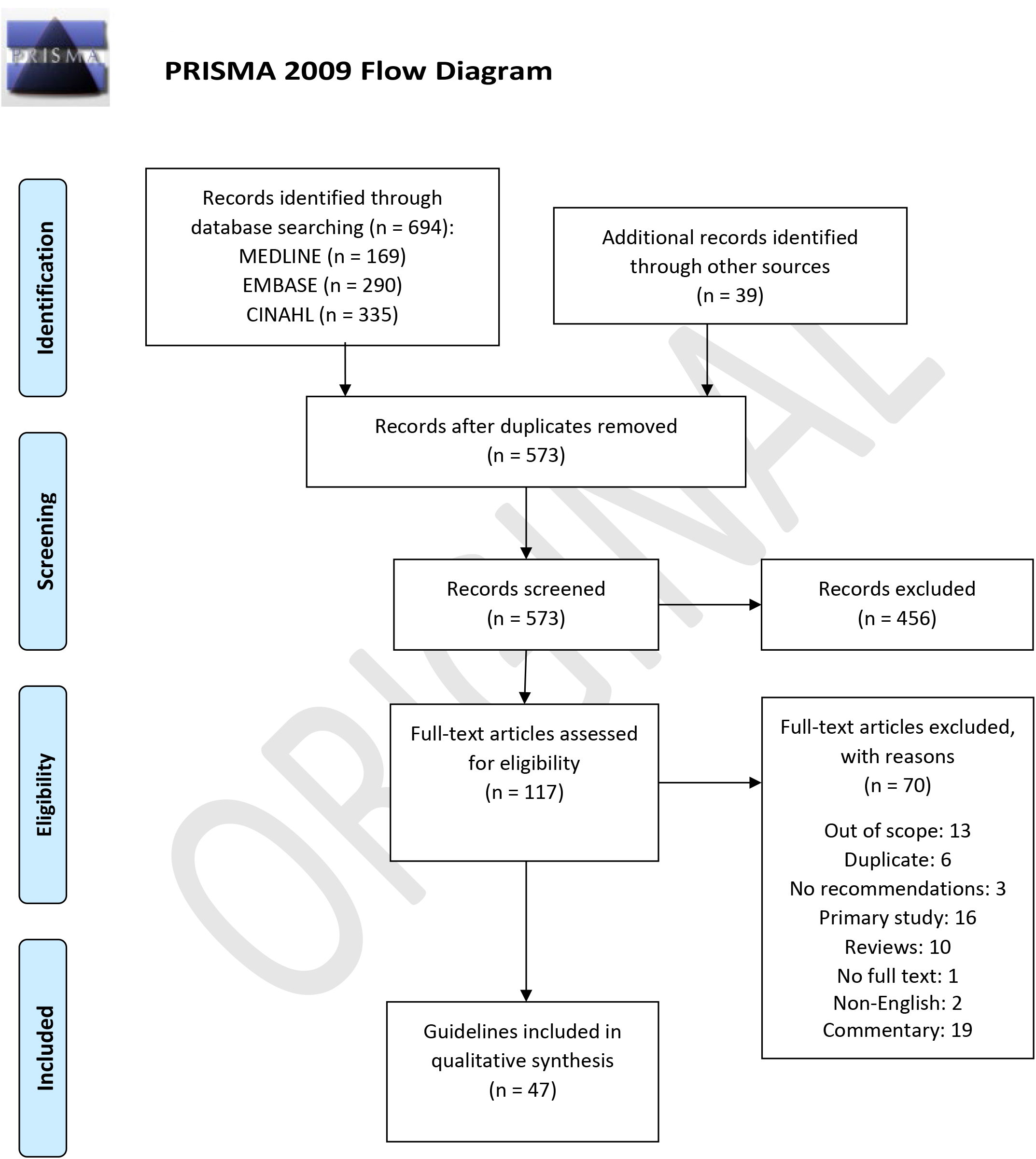
PRISMA to summarize guidelines/policy statements search and selection (https://guides.lib.unc.edu/prisma)

### Characteristics of guidelines

Among the 47 guidelines, 65% were published between 2009 and 2014 (**Table 2**). Most guidelines (35, 74%) targeted adults aged 40 or 50 years and over, and 26% (n=12) were for women only. Based on the World Bank Gross National Income per capita (32), 81% (n=35) were developed in countries at middle-upper or high-income levels, and a majority of the guidelines originated in Europe (16, 34%) and North America (11, 23%). A medical or academic society produced most of the guidelines (42, 89%), and 62% (n=29) did not disclose funding source.

**Table 2.** Characteristics of included guidelines (n=47)

| **Characteristics** | **N** | **Percentage (%)** |
| --- | --- | --- |
| **Year of publication** |  |  |
| 2009-2014 | 31 | 65 |
| 2015-2019 | 16 | 35 |
| **Age distribution** |  |  |
| ≥ 40 years | 13 | 27 |
| ≥ 50 years | 22 | 47 |
| ≥ 60 years | 6 | 13 |
| Adults in general (≥18 years) | 6 | 13 |
| **Sex distribution** |  |  |
| Women | 12 | 26 |
| Men | 1 | 2 |
| Both sexes | 34 | 72 |
| **WHO regions <sup>1</sup>** |  |  |
| Africa | 1 | 2 |
| Americas | 13 | 28 |
| South-East Asia | 2 | 4 |
| Europe | 16 | 34 |
| Eastern Mediterranean | 1 | 2 |
| Western Pacific | 12 | 26 |
| **Other (international guidelines)** | 2 | 4 |
| **Guideline organization category** |  |  |
| Medical or academic society | 42 | 89 |
| Government body | 5 | 11 |
| **Funding source** |  |  |
| None disclosed | 29 | 62 |
| Government agency | 8 | 17 |
| Medical society | 5 | 11 |
| Pharmaceutical / Food Industry | 4 | 9 |
| Mixed sources | 1 | 2 |

<sup>1</sup> World Health Organization. Definition of regional groupings 2020 (45)

### Recommendations on vitamin D and calcium

The daily intake recommendations do not always specify the source intake from diet and/or supplements. For vitamin D, most guidelines (35, 74%) provided a daily intake recommendation, ranging from 200∼600 IU to 4,000 IU per day (**Table 3):** These vitamin D recommendations varied by their sources: 40% (n=19) from diet, 96% (n=45) vitamin D supplements, and 26% (n=12) from outdoor sunlight. For example, a guideline recommended daily intake of 800 IU/day for vitamin D, with recommendations on taking vitamin D supplements at 800 IU/day but not on a specific intake from dietary sources. In another guideline, there were no specific recommendations on daily intake of vitamin D or getting vitamin D from foods, but it recommended taking 400-800 IU/day vitamin D from supplements. Furthermore, 33 guidelines (70%) recommended serum concentration for 25(OH)D, ranging from 25 to 75∼250 nmol/L (i.e., 10 to 30 ∼100 ng/mL), and used words such as “optimal,” “sufficient,” “adequate,” “ideal,” “desirable,” “sustained,” “required,” and “minimal” for the recommended level.

**Table 3.** Percentage of guidelines presenting recommendations on vitamin D and calcium (n = 47)

| Recommendations | Counts | Percentage of guidelines |
| --- | --- | --- |
| **Vitamin D** |  |  |
| **Recommended daily intake (IU/day)** | 35 | 74% |
| 200-600 | 1 | 2% |
| 400-800 | 2 | 4% |
| 400-800; 800-1000<sup>1</sup> | 1 | 2% |
| 400 | 4 | 9% |
| 800 | 6 | 13% |
| 600; 800<sup>1</sup> | 1 | 2% |
| 600 - 800 | 4 | 9% |
| 800 - 1000 | 8 | 17% |
| 800 - 2000 | 2 | 4% |
| >800 | 2 | 4% |
| >1000 | 1 | 2% |
| 1000 - 2000 | 1 | 2% |
| 1500 - 2000 | 1 | 2% |
| 4000 | 1 | 2% |
| **Recommendation on dietary vitamin D** | 19 | 40% |
| **Recommendation on supplemental vitamin D** | 45 | 96% |
| **Recommendation on sunlight exposure** | 12 | 26% |
| **Recommended level of serum 25(OH)D concentration (nmol/L)** | 33 | 70% |
| >25 | 1 | 2% |
| 50 | 3 | 6% |
| 75 | 9 | 19% |
| 80 | 1 | 2% |
| ≥70 | 1 | 2% |
| ≥75 | 3 | 6% |
| >50 | 1 | 2% |
| >60 | 1 | 2% |
| 50-125;75-200<sup>1</sup> | 1 | 2% |
| 50-75 | 4 | 9% |
| 50-80 | 1 | 2% |
| 68-75 | 1 | 2% |
| 75-125 | 4 | 9% |
| 75-150 | 1 | 2% |
| 75-250 | 1 | 2% |
| **Calcium** |  |  |
| **Recommended daily intake (mg/day)** | **33** | **70%** |
| 600 | 1 | 2% |
| 750 | 1 | 2% |
| 700-800 | 1 | 2% |
| 750; 800<sup>1</sup> | 1 | 2% |
| 700-1200 | 1 | 2% |
| 800-1000 | 3 | 6% |
| 1000 | 4 | 9% |
| 1200 | 13 | 28% |
| 1000; 1200<sup>1</sup> | 2 | 4% |
| 1000; 1300<sup>1</sup> | 1 | 2% |
| 1000-1200 | 5 | 11% |
| **Recommendation on dietary calcium** | **34** | **72%** |
| **Recommendation on supplemental calcium** | **38** | **81%** |
| **Recommendation on a whole foods diet to maintain bone health** | **8** | **17%** |

<sup>1</sup> “;” represents separate recommendations exist in the same guideline for different age groups.

For calcium, 33 guidelines (70%) provided daily recommended intake, ranging from 600 to 1,300 mg (Table 3). Most guidelines recommended getting calcium from specific food sources (e.g., dairy) (34, 72%) and supplements (38, 81%).

Regarding recommendations on a whole foods diet, only 17% (n=8) of the guidelines recommended having a nutrition balanced diet to maintain bone health or prevent osteoporosis.

### Assessment of guideline methods

On average, 10 [95% CI of the mean: 9-12; interquartile range: 6-15] of the 25 WHO methodological criteria were met per guideline. Four criteria that were met by more than 70% of the guidelines were: “Priority of the problem stated: Is the problem a burden of disease?” (83%), “Disclosure of conflicts of interest” obtained (74%), “Are recommendations explicitly linked to evidence?” (74%), and “Discipline representation” of the guideline development group (70%) (**Figure 2**). The least frequently met criteria included “Diversity representation” (4%), “Used systematic review methods to synthesize evidence” (11%), “Conflicts of interest managed” (15%), “Outcome importance specified - uncertainty about or variability in how much people value the main outcome?” (19%), “Health equity” considered (21%) and “Acceptability” considered (21%), and external review of guidelines conducted (23%). Detailed descriptions of the WHO guideline development methods criteria are available in **Supplemental Table 1**

**Figure 2.**
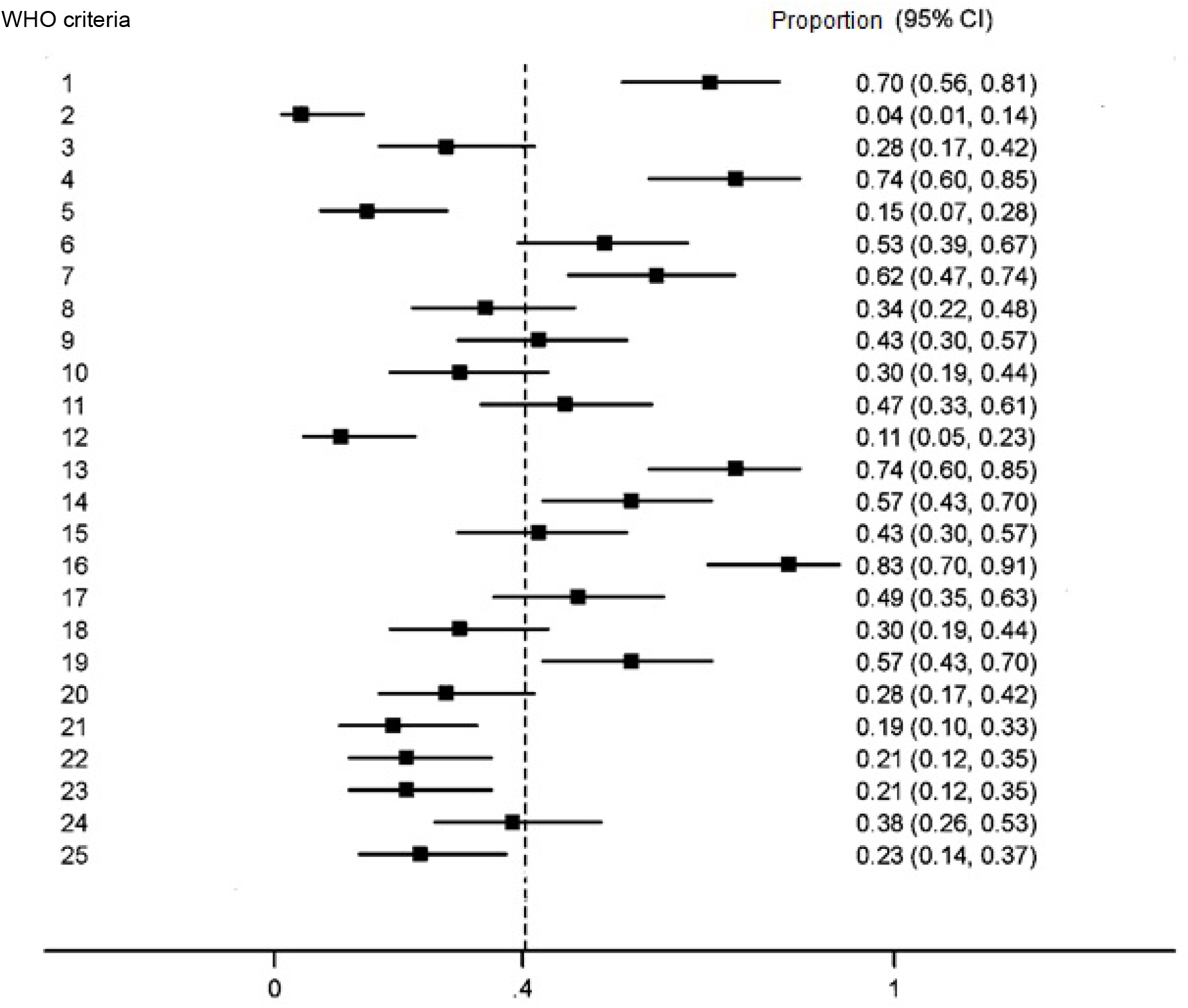
Estimated proportion (95% confidence interval) of guidelines that fulfilled each of the 25 WHO methodological criteria^1^ for guideline development ^**1**^**WHO methodological criteria** (27) : 1. Discipline representation; 2. Diversity representation; 3. Stakeholder input; 4. Disclosure of conflicts of interest obtained; 5. Conflicts of interest managed; 6. Disclosure of funders of the guideline obtained and disclose funder’s role in influencing the guideline development process and recommendations; 7. Formulation of key questions for the evidence review in PICO, PICOT, or PEO format; 8. Choosing (finalizing) priority outcomes for systematic review; 9. Used systematic methods to search for evidence; 10. Used systematic methods to retrieve evidence to select eligible studies; 11. Used systematic methods to assess quality of evidence quality; 12. Used systematic methods to synthesize evidence; 13. Are recommendations explicitly linked to evidence? 14. Was a consensus process clearly described for developing recommendations; 15. Was a method employed to determine strength and/or certainty of the recommendation? 16. Priority of the problem: Is the problem a burden of disease? 17. Quality of the evidence: Is higher quality of the body of evidence included to support the recommendation? 18. Certainty of evidence: Does the recommendation include consistent body of evidence? 19. Benefits and harms: Are evaluations performed on the net benefit or net harm associate with an intervention or exposure? 20. Balance: Does the balance between desirable and undesirable effects support the recommendation? 21. Outcome importance: Is there important uncertainty about or variability in how much people value the main outcome? 22. Equity: Does the evidence used reduce inequalities, improve equity or contribute to the realization of one of several human rights defined under the international legal framework? 23. Acceptability: Is the option acceptable to key stakeholders? 24. Feasibility: Is the option feasible to implement? 25. Was the guideline/recommendation reviewed by an external review group?

There was no clear evidence of an association between the WHO methodological criteria being met and making recommendations for daily intake of vitamin D or calcium (**Table 4**). The CIs for the differences of the proportion of guidelines met a criterion versus those that did not meet the criterion for the relationship of making the recommendations were generally wide, often including potentially important differences at either end of the confidence limits. Even though we found statistically significant results between “Acceptability of recommendations to stakeholders” and recommendation made for daily intake of vitamin D [RD: - 0.38; 99% CI (−0.8, 0.04); p=0.02], and between “guidelines that had undergone an external review” and recommendations made for daily intake of calcium intake [RD: 0.34; 99% CI: 0.14, 0.55; p=0.04], we cannot rule out whether these results were found by chance, due to the small sample size and wide confidence intervals.

**Table 4.**
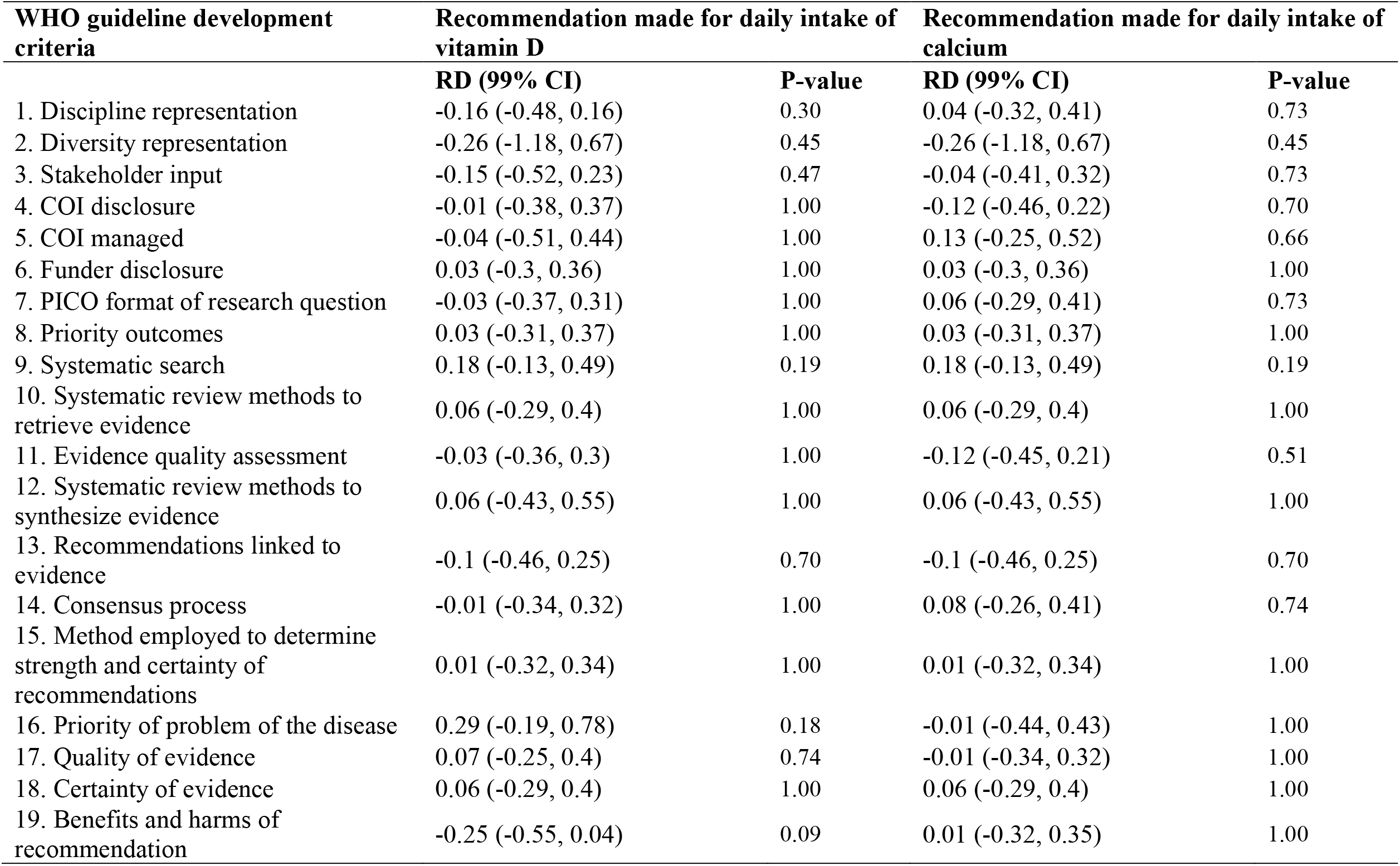
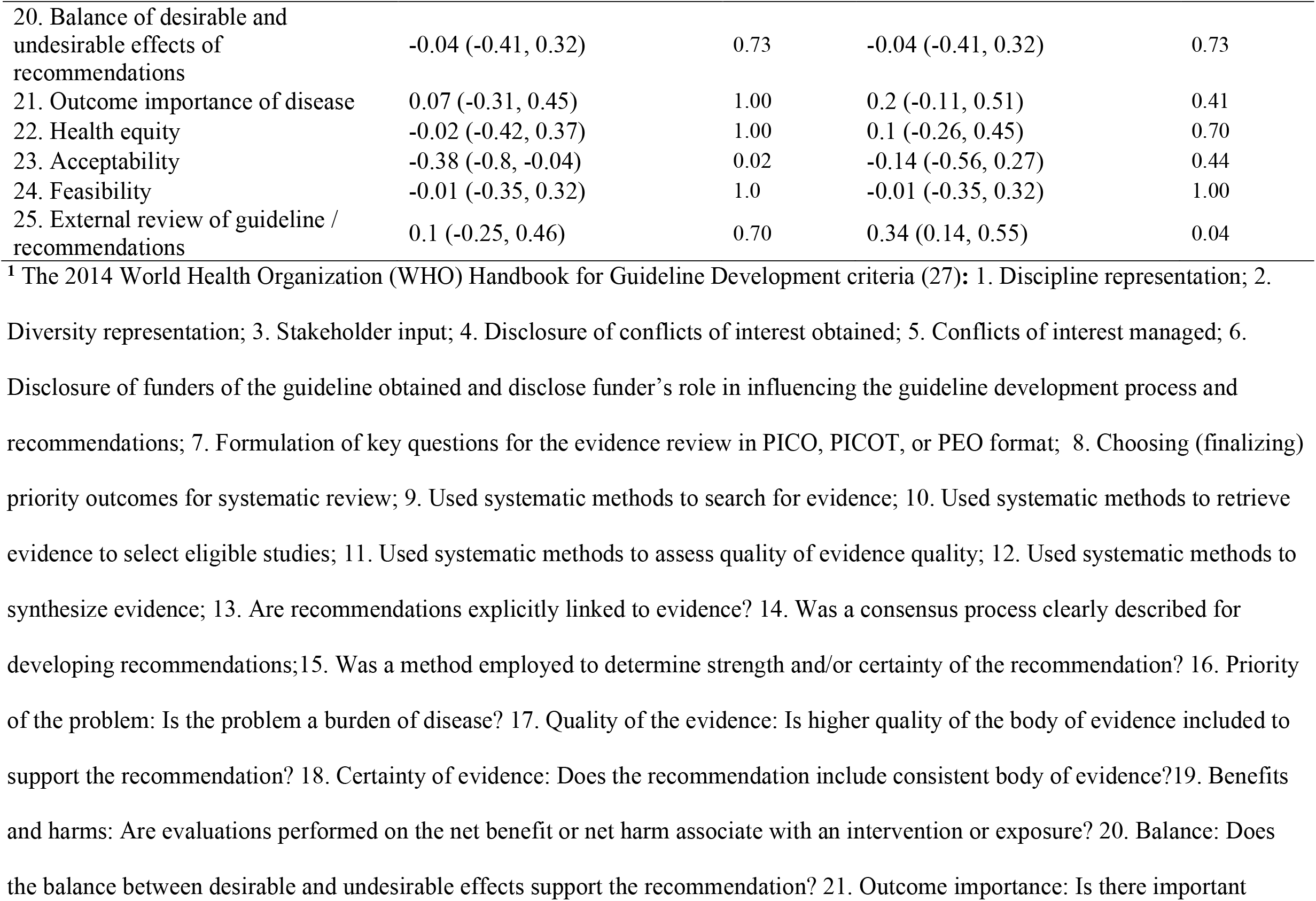
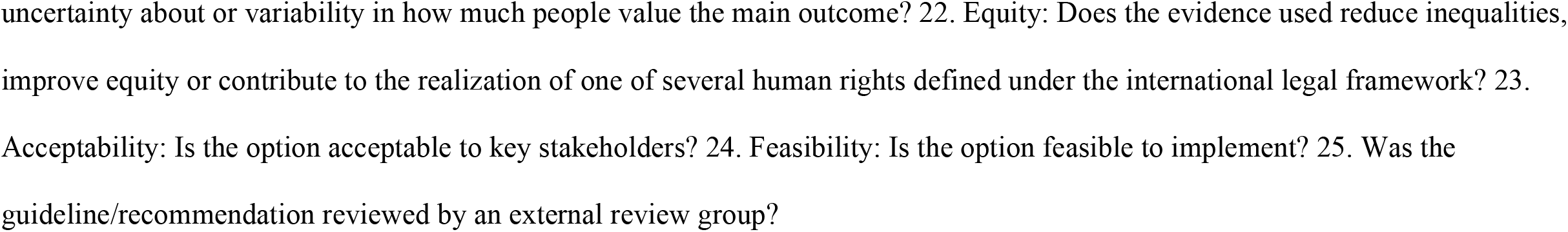
Risk difference (RD) and its 99% confidence interval (CI) for the relationship between a guideline process had met or not met a WHO guideline development criterion^1^ and the recommendations made on vitamin D or calcium

### Evidence cited to support recommendations

Previous guidelines were used most frequently to support the recommendations, except for those on sunlight exposure and supplemental calcium (**Table 5**). For example, 57% (20/35) of the guidelines cited previous guidelines for making recommendations on serum 25(OH)D concentration, and 47% (21/45) on vitamin D supplementation. Among the 49% (18/37; 17/35) that cited previous guidelines on daily intake of vitamin D/calcium, 11 guidelines and 9 guidelines cited government reference values for vitamin D and calcium, respectively, as part of the evidence. Notably, systematic reviews of RCTs were cited in only 8-42% of the guidelines in support of individual recommendations for vitamin D or calcium. Narrative reviews, editorials, or commentaries (33%) constituted the primary supporting evidence for sunlight exposure.

**Table 5.**
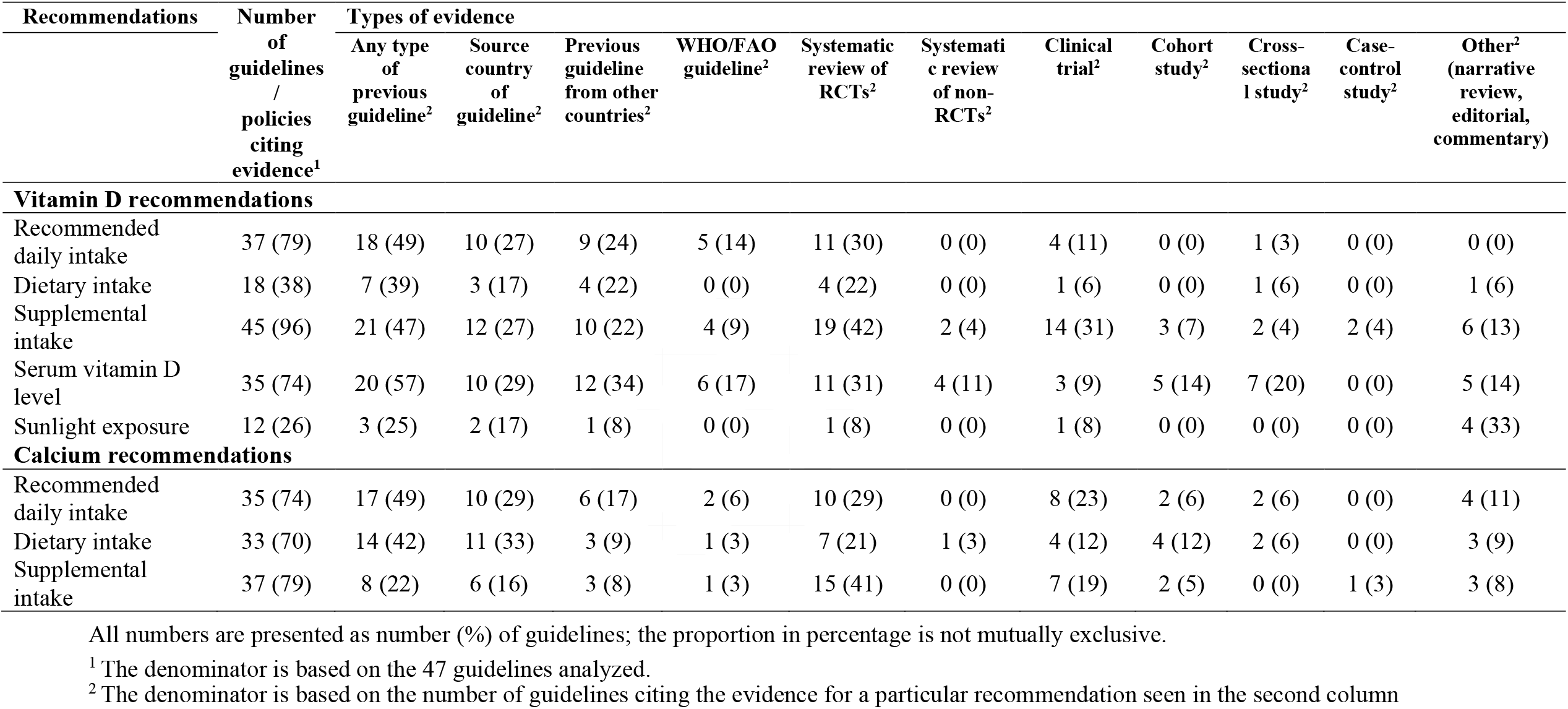
Number and percentage of guidelines (n = 47) citing different types of evidence in supporting the recommendations

| Recommendations | Number of guidelines / policies citing evidence<sup>1</sup> | Types of evidence |  |  |  |  |  |  |  |  |  |  |
| --- | --- | --- | --- | --- | --- | --- | --- | --- | --- | --- | --- | --- |
|  |  | Any type of previous guideline<sup>2</sup> | Source country of guideline<sup>2</sup> | Previous guideline from other countries<sup>2</sup> | WHO/FAO guideline<sup>2</sup> | Systematic review of RCTs<sup>2</sup> | Systematic review of non-RCTs<sup>2</sup> | Clinical trial<sup>2</sup> | Cohort study<sup>2</sup> | Cross-sectional study<sup>2</sup> | Case-control study<sup>2</sup> | Other<sup>2</sup> (narrative review, editorial, commentary) |
| Vitamin D recommendations |  |  |  |  |  |  |  |  |  |  |  |  |
| Recommended daily intake | 37 (79) | 18 (49) | 10 (27) | 9 (24) | 5 (14) | 11 (30) | 0 (0) | 4 (11) | 0 (0) | 1 (3) | 0 (0) | 0 (0) |
| Dietary intake | 18 (38) | 7 (39) | 3 (17) | 4 (22) | 0 (0) | 4 (22) | 0 (0) | 1 (6) | 0 (0) | 1 (6) | 0 (0) | 1 (6) |
| Supplemental intake | 45 (96) | 21 (47) | 12 (27) | 10 (22) | 4 (9) | 19 (42) | 2 (4) | 14 (31) | 3 (7) | 2 (4) | 2 (4) | 6 (13) |
| Serum vitamin D level | 35 (74) | 20 (57) | 10 (29) | 12 (34) | 6 (17) | 11 (31) | 4 (11) | 3 (9) | 5 (14) | 7 (20) | 0 (0) | 5 (14) |
| Sunlight exposure | 12 (26) | 3 (25) | 2 (17) | 1 (8) | 0 (0) | 1 (8) | 0 (0) | 1 (8) | 0 (0) | 0 (0) | 0 (0) | 4 (33) |
| Calcium recommendations |  |  |  |  |  |  |  |  |  |  |  |  |
| Recommended daily intake | 35 (74) | 17 (49) | 10 (29) | 6 (17) | 2 (6) | 10 (29) | 0 (0) | 8 (23) | 2 (6) | 2 (6) | 0 (0) | 4 (11) |
| Dietary intake | 33 (70) | 14 (42) | 11 (33) | 3 (9) | 1 (3) | 7 (21) | 1 (3) | 4 (12) | 4 (12) | 2 (6) | 0 (0) | 3 (9) |
| Supplemental intake | 37 (79) | 8 (22) | 6 (16) | 3 (8) | 1 (3) | 15 (41) | 0 (0) | 7 (19) | 2 (5) | 0 (0) | 1 (3) | 3 (8) |

All numbers are presented as number (%) of guidelines; the proportion in percentage is not mutually exclusive.
<sup>1</sup> The denominator is based on the 47 guidelines analyzed.
<sup>2</sup> The denominator is based on the number of guidelines citing the evidence for a particular recommendation seen in the second column

## Discussion

In this systematic review of 47 global bone health guidelines, we found that the range of recommended daily intake and supplementation of vitamin D and calcium and serum level of 25(OH)D varied substantially; and most of the guidelines recommended taking supplements for bone health and the prevention of osteoporosis. Overall, the methodological quality of these guidelines is low, with, on average, only 10 out of 25 WHO methodological criteria for guideline development being met. Particularly concerning criteria included: lack of diversity of representation in the guideline development group; little description of the management of conflict of interest (COI); limited use of systematic review methods to synthesize evidence; lack of consideration in health equity and acceptability to meet target users’ needs; and there was a paucity of external review of the guidelines. The evidence of the associations between the WHO methodological criteria and variabilities of the recommendations on daily intake of vitamin D or calcium is unclear. Finally, the primary source of evidence underpinning most recommendations was previously published guidelines.

The substantial variability in the recommended levels of vitamin D (200∼600-4000 IU/day) and calcium (600-1200 mg/day), particularly vitamin D, and a lack of systematic reviews as supporting evidence, raises concerns about vitamin D and calcium supplementation recommended in most of the guidelines. As a comparison of daily recommended intakes, the WHO nutrient requirement intake (NRI) for vitamin D is 200 IU/day for adults aged 19–50 years, 400 IU/day for those aged 51–65 years, and 600 IU/day for those aged 65+ years; and 1000 mg/day for 19-65 years / menopause and 1300 mg/day for 65 years+/postmenopause (33). The US Dietary Reference Intakes (DRIs) list the Recommended Daily Allowance (RDA, covering 97.5% of the population) for calcium as 1000 mg/day for men 31-70 years and women 31-50 years and 1200 mg/day for men over 70 years and women over 50 years, and the RDA for vitamin D as 600 IU/day for adults 31-70 years and 800 IU/day for those over 70 years (24). The variations in vitamin D and calcium recommendations in the bone health guidelines in this review, therefore, are not aligned with these government reference values for the recommendations. Moreover, current findings on the effectiveness of vitamin D and calcium supplementation on bone mineral density and on fracture prevention are inconsistent. Several systematic reviews of RCTs have suggested no beneficial effect of vitamin D supplementation on the prevention of total fractures or hip fractures (11-13, 34, 35) or on BMD (11, 12, 14). Two large trials published in 2019 with over 12 months follow-up also found no effects of vitamin D supplementation on bone health (18, 36). Likewise, previous reviews have raised issues on the effectiveness of calcium supplementation on BMD and fracture prevention (10, 37, 38). The effects of vitamin D in combination with calcium supplements on reducing non-vertebral fractures were also inconsistent in systematic review (34, 35). Additionally, adverse events such as hypercalcemia and hypercalciuria (36), fall risks (18), cardiovascular events (16, 19), gastrointestinal symptoms and renal disease (34) have been reported in systematic reviews of clinical trials. Although quite a few of the guidelines in this review were developed more than 5 years ago, and the evidence underpinning the recommendations on vitamin D and calcium was not the most recent, results from several meta-analyses of clinical trials published before and after 2010 have suggested that the effect of vitamin D or calcium supplements on bone mineral density or fracture prevention is uncertain (12, 14, 19, 37), and that the point estimates for supplements of vitamin D with or without calcium for fracture risks did not change materially since before 2010 when these guidelines were released (12). As noted in Table 5, relatively few of the guidelines cited RCTs or systematic reviews of RCTs and we cannot be certain that this was due to availability of RCTs or the methods used by the guideline developers. Along with the low methodological quality of these guidelines, health professionals should be cautious regarding the prescribed daily dose of vitamin D or calcium supplements to individuals at risk of developing osteoporosis or fractures.

With a sample of 47 guidelines and the width of confidence intervals from our results, we were not able to establish whether any of the guideline processes were associated with the recommendations made on daily intake of vitamin D or calcium. Future studies with a large sample size should further examine whether discrepancies in consideration of acceptability of the recommendations, and whether guidelines underwent an external review process would affect decision making on these recommendations. As various physiological factors (6-9) and cultural and religious practices (39) can affect the nutritional needs of vitamin D and calcium in different populations, efforts including increased stakeholder (end users) inputs and an external peer-review process could prompt guideline committees to consider these factors.

Although most guidelines disclosed authors’ COI, only 15% described a procedure to manage it. Disclosure alone does not prevent bias associated with COI; hence any identified COI must be eliminated or managed (40, 41) to reduce the risk of influencing recommendations (42). Likewise, transparency of disclosure and management of COI in bone health guidelines may reduce these potential biases in the recommendations and increase credibility.

We also found that many bone health guidelines used previous bone guidelines to support their recommendations. A similar finding was reported in a review of national dietary guidelines in support of dietary recommendations (43). The high reliance on previous guidelines could be due to the limited capacity of some countries to invest in evidence synthesis for guideline development. However, relying on previous bone health guidelines to support the recommendations may create the risk of perpetuating inadequate guideline development methods and evidence (43). Additionally, the evidence-to-decision frameworks exist, such as the WHO-INTEGRATE framework (44), these were not mentioned as methods used for recommendation formulation.

This review has several strengths. We adopted a comprehensive search strategy to identify global bone health guidelines or policy statements related to vitamin D or calcium recommendations among generally healthy adults aged 40 years and above. Our research questions, search strategy, and planned methods were developed *a priori* and published in a peer-reviewed journal (21). Two reviewers independently appraised guideline development methods using the 2014 WHO Handbook for Guideline Development, a “gold standard” for developing public health and clinical guidelines in the context of global populations. Also, recommendations were independently extracted under standard guidance (26).

This review also has limitations. We primarily relied on what was documented in the published guidelines. This may limit our ability, for criteria marked as “Unclear”, to fully determine whether a process was not implemented or simply not mentioned. Secondly, certain aspects of the development processes were deemed missing if other guideline development standards rather than the WHO methodological criteria were used. Thirdly, although we assessed whether systematic review methods were adopted to identify and evaluate evidence for each guideline, we did not evaluate whether an original systematic review was conducted or whether previous guidelines were the primary source of evidence. Also, we did not assess the methodological rigor of previous guidelines that were used as a primary source of evidence. Including English only publications has limited our sample size and regional coverage, resulting in a higher portion of guidelines from English-speaking countries.

In conclusion, we found that recommendations on vitamin D and calcium vary substantially in a set of 47 bone health guidelines across different countries/regions in the world, and that variability exists in guideline development methods and in types of evidence underpinning the recommendations. Future guideline groups should document efforts to utilize the best available evidence to substantiate nutritional recommendations, implement procedures to eliminate or manage potential conflicts of interest, and address health equity and acceptability in formulating guideline recommendations. This review provides a benchmark of methods and processes used to develop public health guideline recommendations, against which future studies can be compared.

## Data Availability

This is a systematic review including a comprehensive search strategy and references of included and excluded bone health guidelines/policy statements. This information is included in the supplemental materials.

## Acknowledgement

The authors would like to thank Monica Cooper, an Academic Liaison Librarian at the School of Life and Environmental Sciences: Biochemistry, Microbiology and Nutrition Science, Engineering & IT Cluster for her assistance in developing the search strategy in this systematic review. We would also like to thank Dr. Cynthia Kroeger (Charles Perkins Centre, The University of Sydney) for pilot testing the guidelines at the beginning of the project, and Dr. Mark Cooper’s (Professor of Medicine, Head of Clinical School, Concord Clinical School, Faculty of Medicine and Health, University of Sydney) for his constructive comments on the early version of the manuscript.

## Authors’ contributions

Study Design: ZD, LAB;

Data Collection: ZD, SM, and LB;

Methods and Statistics: ZD, JEM, MJP;

Writing: ZD. (first draft);

Revising and editing: All;

Guarantor: ZD, LAB.

All authors have read and approved the final manuscript.

## Abbreviations used

(AGREE) II: Appraisal of Guidelines for Research and Evaluation
NHMRC: the Australian National Health and Medical Research Council
COI: conflict of interest
EAR: estimated average requirement
IU: international unit
RD: risk difference
WHO: World Health Organization
[1, 25(OH)2D3]: 1,25-dihydroxycholecalciferol
[25(OH)D]: 25-hydroxyvitamin-D

